# Association of Left Atrial Structure and Function with Incident Atrial Fibrillation in Black and White Adults: the ARIC Study

**DOI:** 10.64898/2026.07.08.26357526

**Authors:** Yuchen Li, Elsayed Z. Soliman, Srishti Shrestha, Oluseye Ogunmoroti, Faye L. Norby, Daokun Sun, Linzi Li, Amil M. Shah, Lin Yee Chen, Alvaro Alonso

## Abstract

**Background:** Black individuals have a lower incidence of atrial fibrillation (AF) than White individuals despite a higher burden of many traditional cardiovascular risk factors. Differences in left atrial (LA) structure and function by race could partly explain the observed pattern of AF risk.

**Methods:** This analysis included 4,576 (978 Black and 3,598 White) participants from the Atherosclerosis Risk in Communities (ARIC) study, followed between 2011 and 2021. The association of selected echocardiographic measures of LA structure and function with AF incidence was evaluated with race-specific Cox proportional hazards models with adjustment for sociodemographic and clinical covariates. Additional analyses assessed whether LA measures attenuated the association between race and incident AF.

**Results:** The analysis included 778 AF cases (113 in Black and 665 in White participants, mean age 75 years). Larger LA size and worse LA function were associated with higher AF risk in both Black and White individuals, with most associations of similar magnitude in both groups, except for a slightly stronger association of LA reservoir strain in Black than White participants (Black: hazard ratio (HR) 0.89, 95% CI 0.86-0.92 per 1% increase; White: HR 0.94, 95% CI 0.92-0.95, p for interaction = 0.01). In the overall sample, White participants showed higher AF risk compared to Black participants (HR 1.59, 95% CI 1.24-2.03). Adjustment for most individual LA measures did not attenuate the association between race and AF risk.

**Conclusion:** Larger LA size and worse LA function were associated with incident AF in both Black and White ARIC participants. However, these measures did not explain the lower AF incidence observed among Black participants. LA remodeling appears to be an important predictor of AF risk, but it is not the primary explanation for the Black–White AF paradox.

## INTRODUCTION

Atrial fibrillation (AF) is the most common cardiac arrhythmia in adults and is associated with increased morbidity and mortality, including stroke, heart failure, and premature death.^1^ As life expectancy increases and survival from chronic cardiovascular conditions improves with advances in treatment, the burden of AF continues to grow as well.^2, 3^ Although several risk factors of AF are well established, including older age, hypertension, obesity, and underlying cardiovascular diseases (CVD),^4^ these factors do not fully explain differences in AF risk across population groups.

One notable characteristic in AF epidemiology is that Black individuals in the United States experience a lower incidence of AF than White individuals despite a higher burden of many traditional cardiovascular risk factors.^5–9^ Several explanations have been proposed to explain this apparent paradox, including differences in genetic susceptibility correlated with ancestry, socioeconomic and healthcare factors, and potential under-ascertainment of AF among Black individuals.^10–14^ However, none of these factors fully account for the observed racial difference.

Racial differences in left atrial (LA) structure and function may also influence the observed pattern of AF risk.^15^ The left atrium plays a key role in AF occurrence, with LA enlargement and functional impairment contributing to structural and electrical remodeling that facilitate the onset and maintenance of the arrhythmia.^16, 17^ Prior studies have demonstrated that both enlarged LA size and impaired LA function are associated with higher risk of incident AF.^18–21^ However, evidence regarding racial differences in LA structure is inconsistent. Some studies suggest that White individuals have larger LA dimensions after accounting for clinical factors,^22, 23^ whereas others report the opposite.^24^ Additionally, some prior studies have focused primarily on LA size, while few studies involved LA function;^15, 22, 25, 26^ thus, the contribution of LA functional differences to racial differences in AF risk remains unclear.

Thus, we first evaluated associations of multiple echocardiographic measures of LA structure and function with incident AF separately in White and Black individuals. Then, we also examined whether racial differences in LA structure and function explained the association between race and incident AF. By addressing these objectives, this study aims to improve understanding of the potential mechanisms underlying the AF racial paradox and to inform more equitable and targeted strategies for AF prevention.

## METHODS

### Study Population

We conducted a prospective cohort study within the ARIC study, a community-based cohort that enrolled 15,792 adults aged 45 to 64 years from four US communities: Forsyth County (NC), Jackson (MS), suburban Minneapolis (MN), and Washington County (MD), between 1987 and 1989.^27^ Visit 5 (2011-2013) served as the baseline in this study because the detailed echocardiographic assessment was performed at that visit.^28^

Among 15,792 participants examined at Visit 1, 6,538 of them attended Visit 5. We then excluded 754 participants who did not undergo echocardiography at Visit 5, 100 participants who had prevalent AF or missing AF status at Visit 1, 532 participants who had AF diagnosed during the follow-up between Visit 1 and Visit 5, and 544 participants who had any missing covariate data. We also excluded those who were not White or Black from all study field centers and those who were not White from Minneapolis and Washington County due to small sample size (n = 32). The final sample included 4,576 participants (Figure 1). The study protocol was approved by institutional review boards at each field center. All participants provided written informed consent.

**Figure 1:**
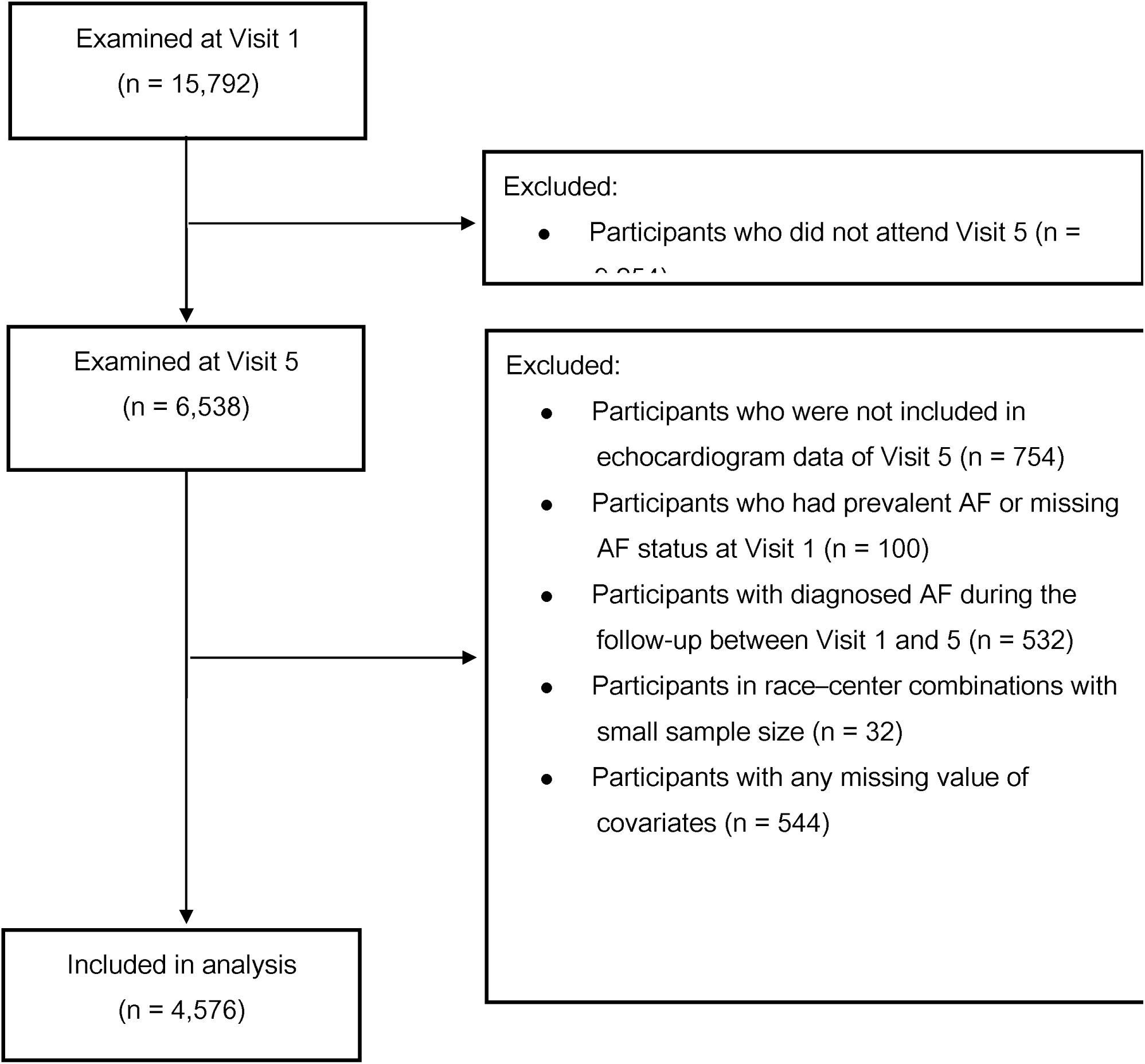
Participants flowchart.

### Echocardiographic Measurements

The exposure of interest included LA size (minimum and maximum LA volume index) and LA function (LA reservoir strain, conduit strain, contraction strain, and emptying fraction). Echocardiograms were performed by dedicated Philips iE33 Ultrasound systems with Vision 2011 and the X5-1 xMatrix transducer for 2D, Doppler, and 3D data acquisition. Images were acquired by trained and certified sonographers. All studies were acquired, stored digitally on a local PACs, and transferred from Field Centers to a secure server at the Echocardiographic Reading Center (Brigham and Women’s Hospital, Boston, MA) same day for central interpretation and quality control. LA strain analysis was performed through QLAB Advanced Quantification Software 13.0 on two-dimensional apical 4-chamber images. LA endocardial borders were traced at end diastole, and phasic LA function was derived from strain- and volume-based measurements over the cardiac cycle. Volume measures were indexed to body surface area.

### AF ascertainment

The primary outcome was incident AF occurring after Visit 5. AF was ascertained based on study electrocardiograms (ECGs), hospital discharge diagnoses, and death certificates. The event date was defined as the first date that a study ECG showed AF, the first hospital discharge with AF diagnosed, or the date of death if AF was recorded in the death certificate, whichever came earlier.^5^ Hospitalization-based events were identified from diagnostic codes of AF or atrial flutter (ICD-9-CM 427.3x or ICD-10-CM I48.x), excluding AF cases occurring in the context of cardiac surgeries (e.g., revascularization) and without evidence of AF in subsequent exams. Follow-up started at the time of Visit 5 until incident AF, death, loss to follow-up, or administrative censoring at December 31, 2021 (through December 31, 2020 at the Jackson site).

### Covariates Measurements

Sociodemographic covariates, including age, sex, race, field center, and education level, were self-reported. Smoking and alcohol consumption status were also ascertained by self-report. Height and weight, used to calculate body mass index (BMI), HDL and LDL cholesterol, systolic and diastolic blood pressure, antihypertensive medication use, and diabetes were obtained by direct measurement, self-report, and from blood samples collected at the time of the visit. Left ventricular mass index and left ventricular ejection fraction from echocardiography were also considered as markers of left ventricular structure and function. Education level was assessed at Visit 1, and other covariates were measured at Visit 5.

Prevalent CHD was defined by self-reported physician diagnosis at Visit 1, evidence of prior myocardial infarction identified from ECGs, or adjudicated CHD events during follow-up. Prevalent stroke at Visit 5 was defined by the criteria from adjudicated cases during follow-up. Prevalent heart failure was defined by multiple sources, including Gothenburg criteria at Visit 1, self-reported use of heart failure medications, or hospitalization records with related diagnostic codes. Diabetes was defined as fasting blood glucose ≥ 126 mg/dL, non-fasting glucose ≥ 200 mg/dL, use of diabetes medication, or self-reported diabetes diagnosis. Hypertension was defined as systolic blood pressure ≥ 140 mm Hg, diastolic blood pressure ≥ 90 mm Hg, or use of antihypertensive medications in past 4 weeks before visit.

### Statistical analysis

Exposures of interest and baseline characteristics were summarized with frequency and percentage for categorical variables and mean and standard deviation for continuous variable.

Initially, we used Cox proportional hazards regression to estimate associations between each LA echocardiographic measure and incident AF separately in Black and White participants. All echocardiographic measures were modeled continuously. Model 1 was a crude model stratified by race; Model 2 was further adjusted for age, sex, center, education level, smoking status, and drinking status; and Model 3 was further adjusted for BMI, prevalent CVD history (CHD, stroke, and heart failure), HDL and LDL cholesterol, systolic and diastolic blood pressure, antihypertensive medication use, diabetes, left ventricular mass index, and left ventricular ejection fraction. To evaluate effect modification by race, we included an interaction term between race and the LA measure of interest in the full cohort models. The proportional hazards assumption was assessed between each variable and time to event.

Then, to evaluate if LA echocardiographic variables accounted for the association between race and AF risk, we fit Cox models with race as the primary exposure and incident AF as the outcome in the entire sample. The base model included both demographic and clinical covariates but not LA echocardiographic measures. Each LA echocardiographic measure was further added separately to assess changes in the hazard ratios comparing risk of AF in Black and White participants accounting for left atrial size and function. Field center was not included in these models because of its collinearity with race in the ARIC recruitment structure. These analyses included participants with non-missing values in all echocardiographic variables (n=3,963) since we compared race effect before and after adding each LA echo markers across models.

Hazard ratios and 95% confidence intervals were reported for all models. A two-sided P-value below 0.05 was considered statistically significant, and an interaction was considered present with a P-value below 0.05 as well. All statistical analyses were conducted by SAS software (version 9.4; SAS Institute Inc., Cary, NC).

## RESULTS

A total of 6,538 participants attended Visit 5, and 4,576 were included in the primary analysis after applying the exclusion criteria shown in Figure 1. Of these, 978 participants were Black and 3,598 were White. Baseline characteristics at Visit 5 are presented in Table 1. The mean age of the analytic cohort was 75.1 years, and 59.1% of participants were female. Compared with White participants, Black participants had a higher mean BMI and a higher prevalence of most cardiovascular risk factors and comorbidities, including stroke, heart failure, diabetes, and use of antihypertensive medications. Black participants also had higher mean systolic and diastolic blood pressure than White participants. In contrast, prevalent CHD was lower among Black than White participants.

**Table 1.** Characteristics of the study population at visit 5 (baseline), stratified by race.

| Sample Characteristic<sup>1</sup> | Total<br>(N=4,576) | Race group |  |
| --- | --- | --- | --- |
|  |  | Black<br>(n=978) | White<br>(n=3,598) |
| **Demographics** |  |  |  |
| Age at baseline, years | 75.1 (5.0) | 74.4 (4.9) | 75.3 (5.0) |
| Sex |  |  |  |
| Female | 2702 (59.1%) | 645 (66.0%) | 2057 (57.2%) |
| Male | 1874 (40.9%) | 333 (34.0%) | 1541 (42.8%) |
| Center |  |  |  |
| Forsyth County, North Carolina | 1022 (22.3%) | 75 (7.7%) | 947 (26.3%) |
| Jackson, Mississippi | 903 (19.7%) | 903 (92.3%) | -- |
| Minneapolis suburbs, Minnesota | 1421 (31.0%) | -- | 1421 (39.5%) |
| Washington County, Maryland | 1230 (26.9%) | -- | 1230 (34.2%) |
| Education Levels |  |  |  |
| Grade school or less | 165 (3.6%) | 91 (9.3%) | 74 (2.1%) |
| High school, but no degree | 408 (8.9%) | 168 (17.2%) | 240 (6.7%) |
| High school graduate | 1531 (33.5%) | 214 (21.9%) | 1317 (36.6%) |
| Vocational school | 396 (8.7%) | 81 (8.3%) | 315 (8.8%) |
| At least some college | 1431 (31.3%) | 216 (22.1%) | 1215 (33.8%) |
| Graduate or Professional school | 645 (14.1%) | 208 (21.3%) | 437 (12.2%) |
| Smoking status |  |  |  |
| Current smoker | 265 (5.8%) | 60 (6.1%) | 205 (5.7%) |
| Former smoker | 2221 (48.5%) | 417 (42.6%) | 1804 (50.1%) |
| Never smoker | 1822 (39.8%) | 394 (40.3%) | 1428 (39.7%) |
| Unknown | 268 (5.9%) | 107 (10.9%) | 161 (4.5%) |
| Drinking Status |  |  |  |
| Current drinker | 2313 (50.6%) | 195 (19.9%) | 2118 (58.9%) |
| Former drinker | 1295 (28.3%) | 435 (44.5%) | 860 (23.9%) |
| Never Drinker | 968 (21.2%) | 348 (35.6%) | 620 (17.2%) |
| **Clinical characteristics** |  |  |  |
| BMI, kg/m<sup>2</sup> | 28.4 (5.3) | 30.1 (6.2) | 27.9 (5.0) |
| Prevalent CHD | 595 (13.0%) | 87 (8.9%) | 508 (14.1%) |
| Prevalent Stroke | 138 (3.0%) | 55 (5.6%) | 83 (2.3%) |
| Prevalent heart failure | 445 (9.7%) | 161 (16.5%) | 284 (7.9%) |
| HDL Cholesterol, mmol/L | 1.4 (0.4) | 1.4 (0.4) | 1.4 (0.4) |
| LDL Cholesterol, mmol/L | 2.7 (0.9) | 2.8 (0.9) | 2.7 (0.9) |
| Systolic blood pressure, mmHg | 130.4 (17.6) | 133.9 (18.8) | 129.4 (17.1) |
| Diastolic blood pressure, mmHg | 66.6 (10.4) | 70.1 (10.3) | 65.6 (10.2) |
| Hypertension Medications in Past 4 Weeks | 2990 (65.3%) | 805 (82.3%) | 2185 (60.7%) |
| Diabetes | 1355 (29.6%) | 387 (39.6%) | 968 (26.9%) |
| **Echocardiogram measures** |  |  |  |
| LA minimum volume index, ml/m<sup>2</sup> | 14.6 (7.4) | 16.0 (8.1) | 14.2 (7.1) |
| LA maximum volume index, ml/m<sup>2</sup> | 25.3 (7.7) | 26.0 (8.1) | 25.2 (7.6) |
| LA reservoir strain, % | 32.6 (7.7) | 31.3 (7.6) | 32.9 (7.7) |
| LA conduit strain, % | -14.8 (5.7) | -14.3 (5.7) | -14.8 (5.6) |
| LA contraction strain, % | -17.8 (5.7) | -16.9 (5.6) | -18.1 (5.7) |
| LA emptying fraction, % | 58.2 (9.4) | 56.4 (9.6) | 58.8 (9.3) |
| LV mass index, g/m<sup>2</sup> | 78.6 (19.5) | 77.5 (21.7) | 79.0 (18.8) |
| LV ejection fraction, % | 65.5 (6.3) | 64.6 (7.0) | 65.8 (6.1) |

<sup>1</sup>Reported as n (%) or mean (SD) unless otherwise specified.
Abbreviations: BMI, body mass index; CHD, coronary heart disease; HDL, high-density lipoprotein; LDL, low-density lipoprotein; LA, left atrial; LV, left ventricular.

Black participants had less favorable LA structural and functional markers than White participants in unadjusted comparisons. Mean LA minimum and maximum volume indices were slightly higher (16.0 vs 14.2 mL/m² and 26.0 vs 25.2 mL/m² respectively) among Black participants, while LA reservoir strain and emptying fraction were slightly lower (31.3% vs 32.9% and 56.4% vs 58.8% respectively). Black participants also had less negative conduit strain and contraction strain values than White participants.

### Associations of LA echocardiographic measures and incident AF

During a mean (median) follow-up of 7.8 (8.9) years, 778 participants were diagnosed with AF (113 Black and 665 White participants). Table 2 shows the associations between LA echocardiographic measures and incident AF stratified by race. Across all three models, associations of LA structure and function were slightly attenuated after adjustment for demographic and clinical covariates, but the overall pattern remained similar. Larger LA size was associated with a higher hazard of incident AF in both race groups. After full adjustment, each 1 mL/m^2^ increase in LA minimum volume index was associated with higher AF risk in both Black participants (1.06, 95% CI 1.03-1.08) and White participants (1.06, 95% CI 1.05-1.07). Similar findings were observed for LA maximum volume index, with HRs of 1.05 (95% CI 1.03-1.07) in Black participants and 1.05 (95% CI 1.04-1.06) in White participants. No significant interaction by race was observed for either LA size measure.

**Table 2:**
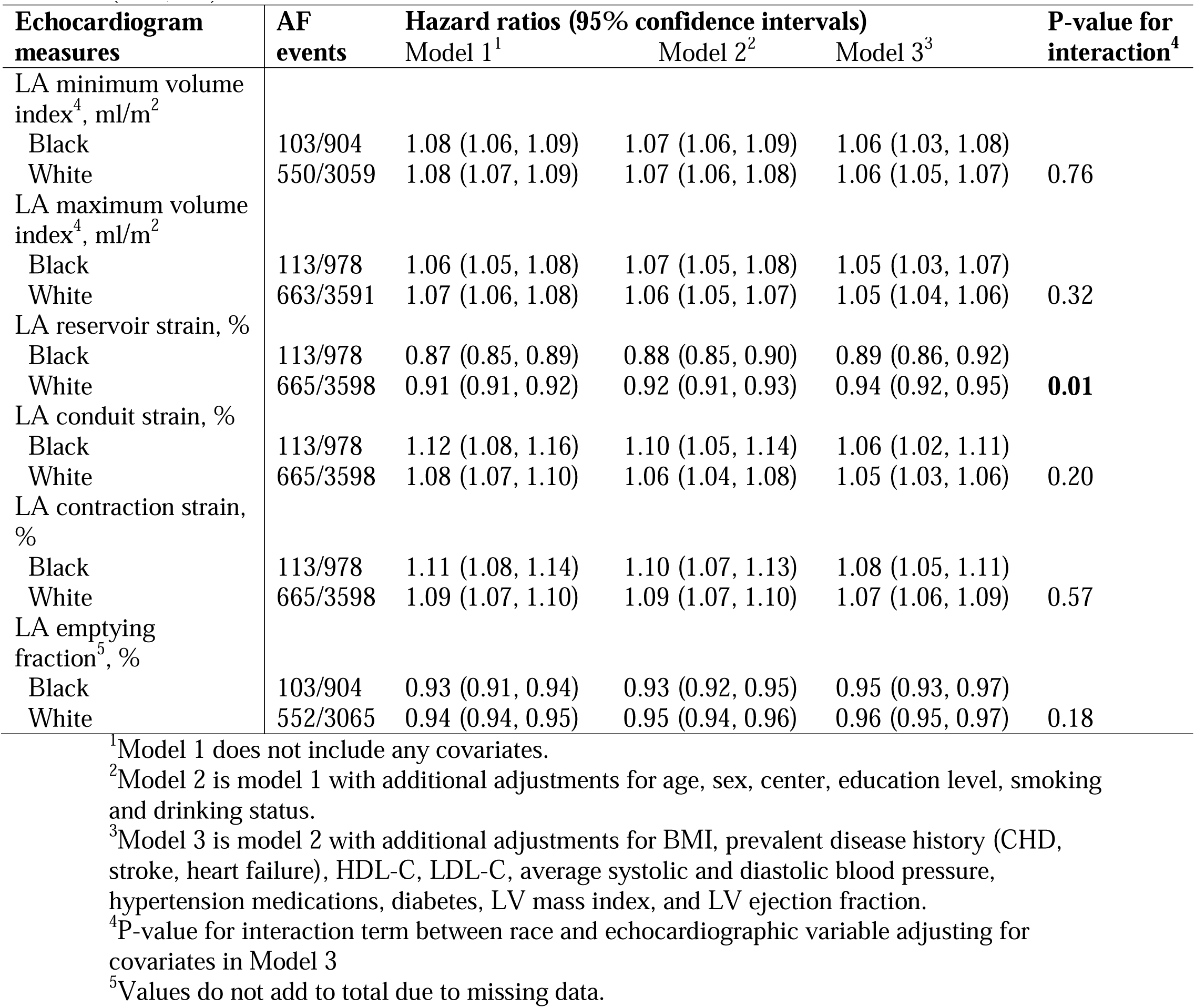
Associations of LA structure and function with incident AF, stratified by race (N=4,576).

LA functional measures were also associated with incident AF in both race groups. Higher LA reservoir strain was associated with lower AF risk in both Black and White participants, and this association was statistically significant. After full adjustment, each 1% higher reservoir strain was associated with a 11% lower AF risk in Black participants and a 6% lower AF risk in White participants. Among all LA measures, only reservoir strain showed statistically significant interaction by race, suggesting that the association between LA reservoir strain and incident AF was stronger among Black participants than White participants (P for interaction = 0.01). LA emptying fraction was also inversely associated with AF in both groups with HRs of 0.95 (95% CI 0.93-0.97) in Black participants and 0.96 (95% CI 0.95-0.97) in White participants, but no significant interaction by race.

LA conduit strain and LA contraction strain were also significantly associated with incident AF in both race groups, but there was not sufficient evidence to prove that these associations differed by race. Overall, Table 2 suggests that adverse LA structure and function were associated with incident AF in both Black and White participants, with only few reductions in the strength of associations between race groups.

### LA measures and racial differences in incident AF

Table 3 shows results evaluating whether LA measures explained the racial difference in incident AF. Among 3,963 participants with non-missing data, White participants had a higher hazard of incident AF than Black participants in the base model (1.59, 95% CI 1.24-2.03; P = 0.0002). After adding each LA echocardiographic measure separately, the HRs comparing White with Black participants (reference) were not attenuated. On the contrary, estimates of association between race and AF risk became stronger after inclusion of each LA marker. For example, additional adjustment for LA volume index increased the HR of AF comparing White to Black individuals to 1.81 (95% CI 1.41-2.32). Similar patterns were observed after adding reservoir strain (1.71, 95% CI 1.34-2.18), contraction strain (1.74, 95% CI 1.36-2.22), and emptying fraction (1.70, 95% CI 1.33-2.17). Adjustment for conduit strain slightly reduced the HR to 1.58 (95% CI 1.23-2.01), which was close to the base estimate and did not suggest meaningful attenuation.

**Table 3:** Contribution of LA Measures to Racial Differences in Incident AF (N=3,963). Table presents the association of race (White vs Black race) with AF risk without adjustment for echocardiographic variables and after including individual echocardiographic variables.

| Echocardiogram variables Added | White race (compared to Black race) |  | LA measure |  |
| --- | --- | --- | --- | --- |
|  | HR (95% CI)<sup>1</sup> | P-value | HR (95% CI) | P-value |
| None (Base Model) | 1.59 (1.24, 2.03) | 0.0002 | -- | -- |
| LA minimum volume index, ml/m<sup>2</sup> | 1.84 (1.44, 2.36) | <.0001 | 1.06 (1.05, 1.07) | <.0001 |
| LA maximum volume index, ml/m<sup>2</sup> | 1.81 (1.41, 2.32) | <.0001 | 1.05 (1.04, 1.06) | <.0001 |
| LA reservoir strain, % | 1.71 (1.34, 2.18) | <.0001 | 0.93 (0.92, 0.94) | <.0001 |
| LA conduit strain, % | 1.58 (1.23, 2.01) | 0.0003 | 1.05 (1.04, 1.07) | <.0001 |
| LA contraction strain, % | 1.74 (1.36, 2.22) | <.0001 | 1.07 (1.05, 1.08) | <.0001 |
| LA emptying fraction, % | 1.70 (1.33, 2.17) | <.0001 | 0.96 (0.95, 0.97) | <.0001 |

<sup>1</sup>Results from Cox proportional hazards model adjusted for age, sex, education level, smoking, drinking status, BMI, prevalent cardiovascular disease (CHD, stroke, heart failure), HDL-C, LDL-C, average systolic and diastolic blood pressure, hypertension medications, diabetes, LV mass index, and LV ejection fraction. Values correspond to hazard ratio (HR) and 95% confidence interval (CI) of atrial fibrillation in White compared to Black individuals.

Overall, these findings do not support the hypothesis that racial differences in LA structure and function explain the lower incidence of AF among Black participants of ARIC cohort, with the Black-White difference in AF risk still present, and even magnified, after adjusting for each LA echocardiographic marker separately.

## DISCUSSION

In this prospective cohort study, larger LA size and worse LA function were associated with incident AF in both Black and White ARIC participants. These associations showed the same trends after adjustment for demographic and clinical covariates, but most interactions with race were not statistically significant. This suggests that the associations between adverse LA structural and functional changes and risk of AF do not differ by race. These findings are consistent with the physiological role of the left atrium, where LA enlargement and impaired function may promote irregular rhythm through structural, electrical and functional remodeling.^18, 19, 29, 30^

Among the LA markers evaluated, only the association between LA reservoir strain and incident AF showed a statistically significant interaction with race. In the fully adjusted model, higher reservoir strain was protective in both groups, but the association appeared stronger among Black participants. Reservoir strain reflects atrial filling and compliance across the cardiac cycle and may capture more integrated impairment in atrial mechanics than volumetric measures alone.^21, 30–32^ Reduced LA reservoir strain may represent a more specific marker of arrhythmogenic atrial myopathy in Black individuals, whereas AF among White participants may arise through a broader range of structural and non-structural mechanisms, explaining the observed interaction. Alternatively, the strongest association in Black participants may reflect differential AF ascertainment if this is affected by LA function, as suggested by studies reporting AF diagnosis at more advanced stages in Black than White participants.^33^ However, this is speculative and requires replication in other cohorts. Thus, the overall findings do not support major racial differences in the associations between most LA markers and incident AF.

In unadjusted analyses, Black participants showed worse LA structural and functional measures at baseline than White participants in this study. However, some earlier studies reported a different pattern. In the Coronary Artery Risk Development in Young Adults (CARDIA) cohort, analyses after full adjustment showed larger LA diameter in White than Black participants, while in patients with ischemic stroke from Greater Cincinnati/Northern Kentucky Stroke Study (GCNKSS) cohort, Black race was also associated with smaller LA diameter after adjustment.^22, 23^ In contrast, prior work in ARIC showed that Black participants had a higher burden of cardiovascular risk factors, greater LA size, and lower LA function than White participants,^24^ which aligned with baseline findings of this analysis. These differences across studies may reflect differences in study population, including age, clinical setting, and the specific atrial phenotype examined. In this study, Black participants showed larger BMI, higher blood pressure, and higher prevalence of heart failure, stroke, and diabetes at the baseline than White participants, which may contribute to the larger LA size among Black participants.

Despite the higher burden of cardiovascular risk factors and worse LA structural and functional measures observed among Black participants, they still showed lower AF risk than White participants in models combining the entire cohort. This pattern is consistent with the Black–White AF paradox. Our results further suggest that those racial difference in LA structure and function do not fully explain the differences in AF risk between Black and White participants in the ARIC cohort. If racial differences in LA structure and function were responsible for the lower AF risk in Black participants, then adjustment for LA markers would have attenuated the race–AF association toward the null. Instead, estimations of HR moved away from the null after adjustment for LA minimum volume index and for LA maximum volume index separately, with similar trends after adjustment for reservoir strain, contraction strain, and emptying fraction. Adjustment for conduit strain only had a minimal change towards the null and did not suggest significant attenuation. Overall, those findings indicate that echocardiographic measures of LA size and function in the analysis do not explain the observed racial difference in AF incidence. Therefore, echocardiographic measures of LA size and function might not be the primary factors of the race disparity in AF incidence.

Other explanations proposed in prior literature,^10–14^ such as differences in genetic susceptibility correlated with ancestry, socioeconomic and healthcare factors, or incomplete AF ascertainment, may still contribute to the paradox. The Black-White AF paradox might be partly driven by genetic ancestry, with higher European ancestry among Black Americans associated with an increased AF risk,^10, 12^ while the single-nucleotide polymorphism rs10824026 could mediate the increased AF risk found in white individuals compared with black individuals.^11^ On the other hand, Black individuals might have lower access to quality care and, therefore, lower ascertainment rates (e.g., less routine screenings in community clinics) may contribute to the paradox.^13, 34^ Thus, the lower AF incidence observed among Black individuals likely reflects a more complex set of mechanisms than differences in the LA markers evaluated in this study.

Several limitations should be considered. First, Black participants in this analysis were recruited from only two ARIC field centers, and the majority were from Jackson, Mississippi. Since race was strongly correlated with field center in the ARIC recruitment, the effects of race and geography cannot be fully separated in the analysis. Only one center (Forsyth County) had both White and Black participants included in the analysis, but the number of Black participants in that center was insufficient to make precise White-Black comparisons. Second, the number of Black participants was smaller than that of White participants, which may result in less precise estimations. Furthermore, AF ascertainment relied on study ECGs, hospitalization codes, and death certificates, which likely misses some asymptomatic or paroxysmal AF events. This could be particularly problematic if underascertainment differs by race or center. However, prior study demonstrated adequate validity of this approach for AF ascertainment in both White and Black ARIC participants.^5^

This study also has several strengths. First, the prospective cohort study design ensured that LA structural and functional measures were assessed before incident AF, which supports the temporality between exposure and outcome. Second, the study included available information on a wide range of potential demographic and clinical confounders, allowing for more precise adjustment in the analysis. Third, the relatively large sample size and number of incident AF events provided adequate power to evaluate associations. Moreover, echocardiographic measurements were obtained using a central reading laboratory, which likely improved the consistency and quality of LA measurements across participants. Additionally, we evaluated both LA size and function measures rather than relying on either of them.

In conclusion, worse LA structure and function were associated with higher risk of incident AF in both racial groups with similar trends. Black participants had relatively worse LA markers at baseline and burden of several cardiovascular risk factors, and adjustment for these markers did not attenuate the racial difference in AF risk. LA echocardiographic markers are associated with incident AF but are not the primary explanation for the race paradox in AF incidence. Further research is needed to identify and confirm other reasons for this racial difference.

## Data Availability

All data produced are available upon reasonable request to the ARIC Coordinating Center.

## Nonstandard Abbreviations and Acronyms

AF: atrial fibrillation
ARIC: Atherosclerosis Risk in Communities
CVD: cardiovascular diseases
LA: left atrial

## ACKNOWLEDGMENTS

The authors thank the staff and participants of the ARIC study for their important contributions.

